# An urgent need for a paradigm shift in HIV testing for older children: A sine qua non condition to achieve an AIDS-free generation

**DOI:** 10.1101/2020.06.22.20128363

**Authors:** H.A. Yumo, J.J.N. Ndenkeh, I. Sieleunou, D.N. Nsame, P.B. Kuwoh, M. Beissner, C. Kuaban

## Abstract

**Background:** Achieving an AIDS-free generation requires effective pediatric testing and treatment services. While pediatric HIV testing has been more focused on children below 18 months through PMTCT, the yield of this approach remains unclear comparatively to testing children above 18 months through routine PITC. This study aimed at bridging this evidence gap and provide knowledge to guide pediatric HIV testing investments.

**Materials and Methods:** Parents visiting or receiving HIV care in three hospitals in Cameroon were invited to test their children for HIV. HIV testing was done using PCR and antibody rapid tests for children < 18 months and those ≥18 months, respectively. We compared HIV case detection and ART initiation between the two subgroups of children and this using Chi-square test at 5% significant level.

**Results:** A total of 4079 children aged 6 weeks-15 years were included in the analysis. Compared to children < 18 months, children group ≥18 months was 4-fold higher among those who enrolled in the study (80.3% vs 19.7%, p<0.001); 3.5-fold higher among those who tested for HIV (77.6% vs 22.4%, p<0.001); 6-fold higher among those who tested HIV+ (85.7% vs 14.3%, p=0.241) and 11-fold higher among those who enrolled on ART (91.7% vs 8.3%, p< 0.028).

**Conclusions:** Our results show that 4 out of 5 children who tested HIV+ and over 90% of ART enrolled cases were children ≥ 18 months. Thus, while rolling out PCR HIV testing technology for neonates and infants, committing adequate and proportionate resources in antibody rapid testing for older children is a sine quo none condition to achieve an AIDS-free generation.

## Introduction

The HIV/AIDS pandemic remains a global health threat. According to the UNAIDS estimates, 37.9 million people were living with HIV/AIDS globally in 2018, including 1.7 million children (<15 years). In that same year, 770 000 persons died of HIV/AIDS in the world, including 100 000 children. Africa hosts 68% of PLHIV worldwide and in 2018, 62% of AIDS related deaths were from this continent (1). Despite this burden, transformative and remarkable progress has been made in the past decades with regards to the expansion of antiretroviral therapy (ART)(2–4). In 2018, there were 23 300 000 PLHIV receiving ART, representing 62% of patients in need of treatment. At global level, this treatment expansion has contributed in the reduction of HIV incidence and mortality by 16% and 33%, respectively, from 2010 to 2018(1). However, children are not benefiting enough from this treatment expansion as there were 54% children on ART against 62% of adults as of June 2019(5). Cameroon (West and central Africa) with an HIV prevalence of 3.6% has 540 000 PLVIH, including 43 000 children. In 2018, 14 000 people died of AIDS in this country, including 3 600 children. The gap in pediatric ART is even wider in Cameroon as 24% of eligible children are on treatment against 55% of adults (6).

Multiple factors account for current gaps in pediatric and adolescent ART coverage. However, poorly-implemented and/or ineffective case-finding strategies are major barriers to ART roll out in this sub-population (7–9). More importantly, though achieving an AIDS-free generation requires the effective implementation of both prevention and treatment HIV services(10), the global agenda in the past decades has been focused more but on prevention. In this regard, the prevention of mother to children transmission of HIV (PMTCT) program has received more attention and resources from donors, policymakers, practitioners(11). Though the fourth prong of PMTCT include the provision of treatment to HIV-infected infants, this program provides services to infants only from birth up to 18 months(12). Consequently, HIV testing and treatment services for older children (≥18 months) has received less attention in the past decades. Actually, out of the PMTCT program, older children (≥18 months) are expected to be screened for HIV in health facilities through the WHO routine provider-initiated counseling and testing (PITC) (13). Unfortunately, PITC implementation has been suboptimal across countries and this for several reasons including fear of stigma among parents/caregivers, lack of staff training, lack of HIV testing kits, poor commitment from facility leadership, and missed parental consent to test children(14–16). These weaknesses have left many HIV infected children undiagnosed and untreated in the community to the extent that pediatric HIV has been considered as neglected disease by some authors (17).

The target population for pediatric HIV infection are children from 0-15 years(1). Thus, achieving an AIDS free generation requires investing adequate resources in testing and treatment across this age group. As per WHO guidelines, children less than 18 months (in PMTCT) are HIV tested using polymerase chain reaction (PCR) while those older than 18 months (in PITC) are tested using antibody rapid tests(18). While more focus and emphasis have been on PCR testing, the yield of this approach in terms of pediatric HIV case detection and ART initiation compared to antibody rapid tests remain unclear. This study intends to bridge this scientific gap and provide information needed to guide pediatric HIV testing investments. This should contribute in reducing the current gap in HIV treatment for children in Cameroon and beyond.

## Methods

### Design

This was a secondary analysis of the dataset of the “Active Search for Pediatric HIV/AIDS” (ASPA) project published elsewhere (19). ASPA was a larger study which compared the acceptability, feasibility and effectiveness of targeted provider-initiated testing and counseling (tPITC) versus blanket provider-initiated testing and counselling (bPITC) in Cameroon. In this study, we invited all HIV infected parents receiving HIV care in three hospitals in Cameroon to have their children of unknown HIV status aged 6 weeks to 19 years tested for HIV (tPITC group). In the same hospitals, all parents/guardians who accompanied their sick children of the same age group for consultation at the outpatient departments were also counseled, and these children were invited to test for HIV irrespective of the presenting complaint (bPITC group).

### Setting

The ASPA study was implemented at three public health facilities: Limbe Regional Hospital (LRH) in the South-West, Ndop District Hospital (NDH) in the North-West and Abong-Mbang District Hospital (ADH) in the Eastern Region. These facilities provide comprehensive healthcare services, including HIV testing and treatment and were purposefully selected for inclusion of urban, semi-urban and rural populations.

### Study period and population

The ASPA study was implemented at LRH from July through December 2015, and from June through November 2016 at the two other sites. The study population in the tPITC group consisted of parents living with HIV/AIDS receiving care in the hospital and their children of unknown HIV status, aged 6 weeks to 19 years. Similarly, in the bPITC group, the study population consisted of parents/guardians and their sick children of the same age group who attended the hospital outpatient department for any reason. Children or parents critically ill (in vital distress) were excluded from the study.

### Study procedures

#### Enrollment of participants and data collection

In the tPITC group, HIV-positive parents in care at the HIV treatment center (ART clinic) were counseled and invited by a trained counselor to participate in the study together with their children. These parents were offered a testing opportunity for their biological children in either the hospital or at home (community testing). In the bPITC group, parents/guardians were also counseled and invited to have their sick children tested for HIV irrespective of the reason of consultation.

In both groups, all parents/guardians who consented to participate in the study were enrolled together with their children. Pre-tested and structured questionnaires were used by a trained data clerk to collect socio-demographic information of participants. For children, we collected information on age, sex, educational level and HIV testing history (previous HIV testing and result).

#### HIV testing and ART enrolment

For children younger than 18 months of age, HIV testing was performed using DNA-PCR techniques. For children aged 18 months and above, HIV testing was performed using two HIV antibody rapid tests according to the Cameroon national guidelines. During the study period, the WHO test and treat policy was not yet effective at the site level. Thus, children who tested HIV positive were assessed for ART eligibility using WHO clinical staging and/or baseline biological analysis, including CD4 count. Eligible children were initiated on ART and monitored according to the Cameroon national guidelines.

#### Data management and analysis

Anonymous data from the questionnaires were entered into a database and analyzed using STATA 2013 (College Station, TX: StataCorp LP). For the purpose of this article, we excluded from the ASPA dataset, all parents as well as children 15 years or above. Only children less than 15 years were included in the analysis and this because this is the age limit for paediatric HIV/AIDS. The study outcomes were determined by computing and comparing the proportions using Chi-square or Fisher’s exact tests at 5% significant level.

#### Ethical considerations

Participation in the study was voluntary for both parents and children. Only parents who consented were enrolled and assent was requested from children above 11 years of age. Consent from parents was obtained via signed written consent form. Likewise, assent for children over the age of 11 years was obtained through a signed written assent form. The ASPA study received ethical approval from the Cameroon National Ethics Committee, the Ludwig-Maximilians-Universität, Munich (Germany) and the Albert Einstein College of Medicine (NY, U.S.). The ASPA study was also registered at clinicaltrial.gov (NCT03024762) (19). This study was also approved by the Cameroon Ministry of Public Health.

## Results

A total of 4719 children and adolescents were enrolled in the ASPA study. 640 adolescents who were 15 years of age or above were excluded from the dataset. Among children/adolescents included in this analysis (n=4079), the age ranged from 6 weeks to 14 years, 51.8% were males, 49.6% had primary school level, 71.1% enrolled in the study through their mothers, and only 24.8% had previously tested for HIV (Table 1).

**Table 1:**
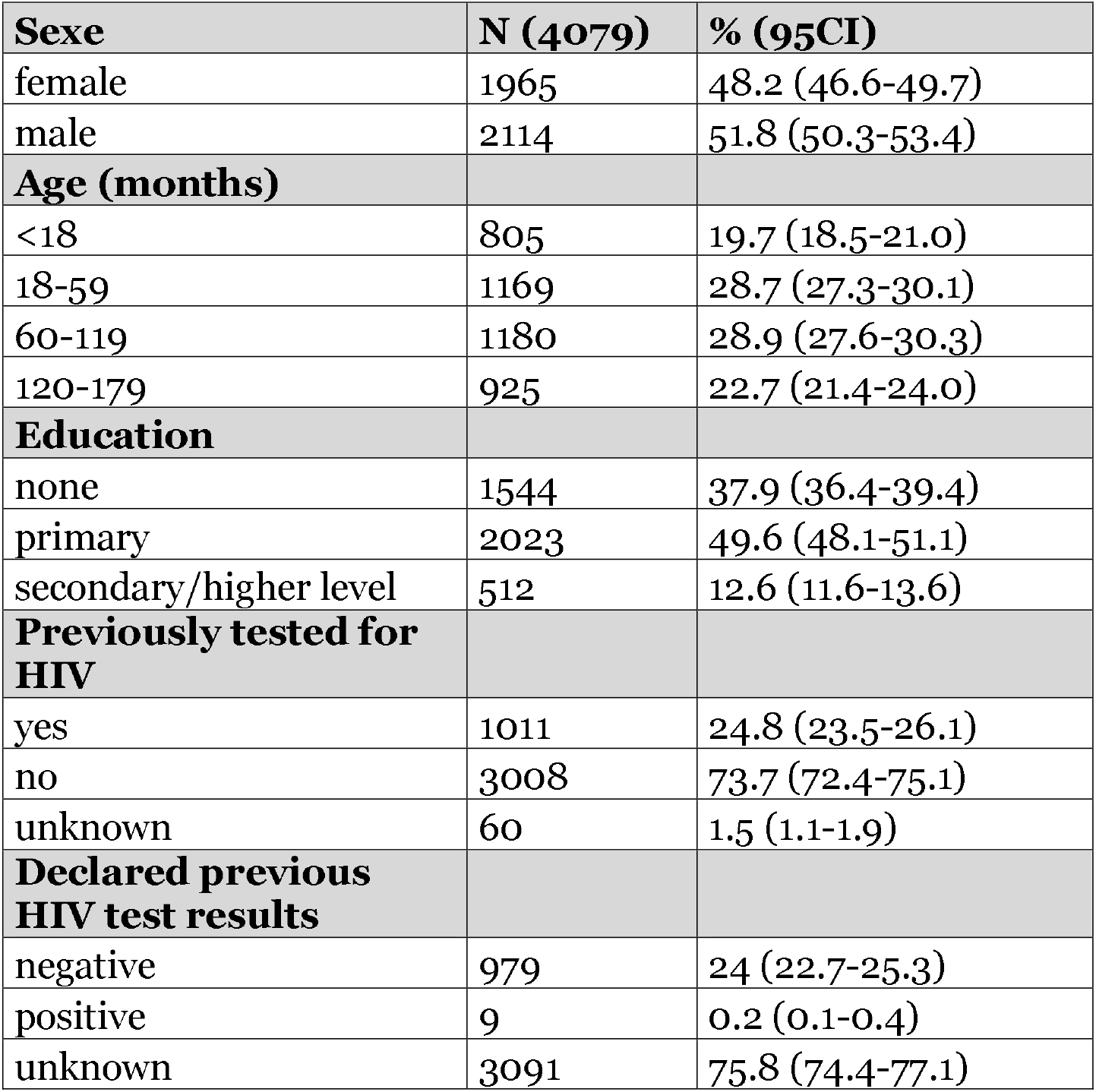
Characteristics of children and adolescents enrolled in the study

|  |  |  |
| --- | --- | --- |
| **Sexe** | **N (4079)** | **% (95CI)** |
| female | 1965 | 48.2 (46.6-49.7) |
| male | 2114 | 51.8 (50.3-53.4) |
| **Age (months)** |  |  |
| <18 | 805 | 19.7 (18.5-21.0) |
| 18-59 | 1169 | 28.7 (27.3-30.1) |
| 60-119 | 1180 | 28.9 (27.6-30.3) |
| 120-179 | 925 | 22.7 (21.4-24.0) |
| **Education** |  |  |
| none | 1544 | 37.9 (36.4-39.4) |
| primary | 2023 | 49.6 (48.1-51.1) |
| secondary/higher level | 512 | 12.6 (11.6-13.6) |
| **Previously tested for HIV** |  |  |
| yes | 1011 | 24.8 (23.5-26.1) |
| no | 3008 | 73.7 (72.4-75.1) |
| unknown | 60 | 1.5 (1.1-1.9) |
| **Declared previous HIV test results** |  |  |
| negative | 979 | 24 (22.7-25.3) |
| positive | 9 | 0.2 (0.1-0.4) |
| unknown | 3091 | 75.8 (74.4-77.1) |

The age disaggregation shows that 19.7% and 80.3% of enrolled children/adolescents were < 18 and ≥18 months of age, respectively. These proportions correspond to children /adolescents eligible for PCR HIV testing (<18 months) and HIV serology (≥18 months), respectively. Overall, 3156 (77.4%) children/adolescents were tested for HIV. Of those, 22.4% and 77.6% were < 18 months and ≥18 months, respectively. This difference was statistically significant (p<0.001). Among children/adolescents who tested for HIV, 98.1% (3095) received their results. Among those who did not receive their HIV test result (n=61), 90.2% and 9.8% were children/adolescents < 18 months and ≥18 months, respectively. This difference was statistically significant (p<0.001). Among children/adolescents who tested for HIV, 63 (2.0%) were found HIV+. Of those, 9 (14.3%) and 54(85.7%) were children < 18 and ≥18 months, respectively. This difference was not statistically significant (p= 0.241). Among children/adolescents who tested HIV+, 48 (76.1%) were enrolled on ART. Children/adolescents < 18 months and those ≥18 months contributed by 8.3% and 91.7% (p< 0.028), respectively to the total number of children/adolescents who enrolled on ART (Table 2).

**Table 2:** Contribution of children’s age in HIV testing, case detection and ART enrolment

|  |  |  |  |  |  |
| --- | --- | --- | --- | --- | --- |
| Outcomes |  | Total<br>n (col %) | age <18<br>months<br>n (row%) | age ≥ 18<br>months)<br>n (row%) | p value* |
| Children eligible for rapid antibody test | No | 805 (19.7) | 805 (19.7) | 0 | N/A |
|  | Yes | 3274 (80.3) | 0 | 3274 (80.3) |  |
| Children tested | No | 923 (22.6) | 99 (10.7) | 824 (89.3) | <0.001 |
|  | Yes | 3156 (77.4) | 706 (22.4) | 2450 (77.6) |  |
| Results received | No | 61 (1.9) | 55 (90.2) | 6 (9.8) | <0.001 |
|  | Yes | 3095 (98.1) | 651 (21.0) | 2444 (79.0) |  |
| Children’s HIV results | Negative | 3032 (98.0) | 642 (21.2) | 2390 (78.8) | 0.2413 |
|  | Positive | 63 (2.0) | 9 (14.3) | 54 (85.7) |  |
| Children enrolled on | No | 15 (23.8) | 5 (33.3) | 10 (66.7) | 0.028 |
|  | Yes | 48 (76.1) | 4 (8.3) | 44 (91.7) |  |
| ART |  |  |  |  |  |
| P value*= Chi-square or Fisher’s exact test |  |  |  |  |  |

## Discussion

According to WHO, “efforts in the global research agenda on pediatric HIV must be focused on generating targeted evidence that improves HIV programme implementation through a better understanding of what works for infants and children”(20). Our study is a response to this need.

We found that the HIV testing uptake was statistically higher among children > 18 months compared to those < 18 months (77.6% vs 22.4%, p<0.001). Indeed, among children enrolled in the study, those ≥18 months of age were 3-fold more likely to be tested for HIV compared to those < 18 months. This was due to the fact that during the study period, the HIV antibody rapid tests were readily available in the laboratory, unlike the DBS-PCR testing kits that were affected by the recurrent stock outs during that same period. The stock outs of PCR testing commodities had been previously reported as factors affecting the early infant diagnosis (EID) of HIV among neonates in sub-Saharan Africa countries(21,22). The use of point-of-care (PoC) HIV testing technology could address this gap and improve the turn-around time of EID results as well as the early ART initiation among HIV infected neonates(23–25).

Among children who tested for HIV, 98.1% received their results. Of those who did not receive results, the proportion of children < 18 months were significantly higher compared to children ≥18 months (90.2% vs 9.8%, p<0.001). This was mainly due to the long turned around time of EID PCR testing for children < 18 months and the indeterminate HIV results for those ≥18 months. The use of PoC technology as indicated above and the availability of 3^rd^ HIV rapid test (tie-breaker) in the laboratory should improve this outcome.

Among children who tested for HIV (n=3156), 63 (2%) tested HIV+. Among those children ≥ 18 months were predominantly represented compared to those < 18 months (85.7% vs 14.3%, p=0.2413). Though this difference was not statistically significant, the HIV positivity among children ≥ 18 months was 6-fold higher compared to that of children < 18 months. This suggests that a greater proportion of children living with HIV/AIDS are found among children ≥18 months. According to UNAIDS estimates, our finding is in line with the proportion of children < 18 months living with HIV/AIDS in Cameroon and elsewhere (26). Therefore, adequate resources should be invested in testing children ≥ 18 months using HIV rapid tests.

Among children tested HIV+ (n=63), 76.1% were enrolled on ART. Of those, the proportion of children ≥ 18 months was significantly higher compared to those < 18 months (91.7% vs 8.3%, p= 0.028). Actually, the contribution of children ≥18 months in the paediatric ART cohort was 11-fold higher compared to children < 18 months. This further demonstrates the need of adequate investments in case finding strategies among the subpopulation of children ≥ 18 months in Cameroon and probably beyond.

## Policy implications

Our results show that children ≥ 18 months have the highest contribution with regards to paediatric HIV case load and ART enrolment. This indicates the role the rapid HIV test could play in bridging the current gap in paediatric HIV care. That notwithstanding, HIV testing for older children is still limited in most high burden paediatric HIV countries. In these countries, the most common entry point for HIV testing of children ≥ 18 months is the outpatient paediatric consultations or inpatient paediatric wards using the WHO recommended PITC(13). However, as indicated above PITC implementation had remained suboptimal across countries. As a result, only approximately 10% and 15% of HIV-infected young (15-24 years) males and females, respectively, in Sub-Saharan Africa are aware of their HIV status (27). This is a clear indication that many children ≥ 18 months are not adequately tested for HIV despite the higher HIV seropositivity among them as indicated by this study. This finding urges for a paradigm shift to give HIV testing for older children the adequate and proportionate attention and resources. This is a sine qua non condition to achieve the AIDS-free generation in high burden paediatric HIV countries.

We believe that in addition to all the barriers to the uptake of PITC reported to date(28,29) the lack of adequate indicators to monitor the implementation of this strategy at facility level is a significant contributing factor to the current low uptake of PITC at facility level. Indeed, to monitor the performance of health facilities regarding HIV testing of their clients, it is important for these facilities to report both the number of outpatient consultations (denominator) and the number of patients tested for HIV (numerator). However, unlike the numerator, the denominator is most often not captured in the monthly summary HIV testing reports. This results to the lack of routine monitoring of health facilities performance with regards to HIV testing strategies, including PITC. On the other hand, other entry points for testing of older children (≥18 months) had not been adequately used by care providers for paediatric HIV case finding across countries in sub-Saharan Africa, including Cameroon. For example, though recommended by WHO since 2010(30), the uptake of index case HIV testing, using HIV infected parents in contact with health facilities (e.g: ART clinics) remains low in these countries. Recent studies from Kenya (31), Malawi (32) and Cameroon (19,33,34) have demonstrated that index case testing is a high-yield strategy for paediatric HIV case finding. Thus, in addition of monitoring the denominator of clients visiting the health facilities, the adequate implementation of index case testing among children ≥ 18 months should also contribute significantly in improving ART expansion among children.

## Strengths and limitations

To the best of our knowledge, this is the first analysis from primary data examining the contribution of the PCR HIV testing technology against the rapid HIV antibody tests in paediatric HIV case detection and ART enrolment. Thus, our study has filled an important scientific gap in ART expansion among children. However, our study is limited by the fact that the sites were not randomly selected and therefore, our results may have been affected by selection bias. That notwithstanding, the external validity of our study is stronger as it was conducted in 3 hospitals from three different geographic locations covering urban, semi-urban and rural populations in Cameroon. Hence, this study has provided new evidence that should guide the investments of the already scarce HIV funding in achieving greater impact in resource-limited settings.

## Conclusions and recommendations

This study shows that 4 out of 5 children found HIV+ and over 90% of those enrolled on ART were children ≥ 18 months. Therefore, while rolling out the POC HIV testing technology for children below 18 months, committing adequate and proportionate resources in testing children above 18 months should be a smart investment. Translating this evidence into practice would require tweaking the data collection tools so as to ensure the routine monitoring of HIV testing performance of health facilities, but also expanding HIV testing to all high-yield entry points. These adjustments are necessary to reducing the current gap in paediatric HIV treatment, and to achieve the AIDS-free generation in Cameroon and beyond.

## Data Availability

The dataset of this paper is available upon request.

## Authors’ Contributions

**HAY:** conceived, conceptualized and designed the study, drafted the study protocol, fundraised for the study, recruited and trained study staff, supervised data acquisition, analyzed data, interpreted the results and drafted the manuscript.

**JJNN:** collected data, supported data analysis, reviewed the manuscript.

**IS:** co-supervised data acquisition, interpreted the results and reviewed the manuscript.

**DNN:** co-supervised data acquisition and reviewed the manuscript.

**PBK:** co-supervised data acquisition, interpreted the results and reviewed the manuscript.

**MB:** Reviewed the study protocol, interpreted the results and reviewed the manuscript. CK: Reviewed the study protocol, interpreted the results and reviewed the manuscript.

## Acknowledgements

The authors are thankful to the study participants and the research assistants.

## References

1. UNAIDS. UNAIDS Data 2019 [Internet]. Geneva, Switzerland; 2019. Available from: https://www.unaids.org/sites/default/files/media_asset/2019-UNAIDS-data_en.pdf

2. Teasdale CA, Abrams EJ, Yuengling KA, Lamb MR, Wang C, Vitale M, et al. Expansion and scale-up of HIV care and treatment services in four countries over ten years. PLOS ONE. 2020 Apr 16;15(4):e0231667.

3. Ford N, Calmy A, Mills EJ. The first decade of antiretroviral therapy in Africa. Glob Health. 2011 Sep 29;7:33.

4. El-Sadr WM, Harripersaud K, Rabkin M. Reaching global HIV/AIDS goals: What got us here, won’t get us there. PLoS Med [Internet]. 2017 Nov 7 [cited 2020 May 15];14(11). Available from: https://www.ncbi.nlm.nih.gov/pmc/articles/PMC5675304/

5. UNAIDS. FACT SHEET – WORLD AIDS DAY 2019 [Internet]. Geneva, Switzerland; 2019. Available from: https://www.unaids.org/sites/default/files/media_asset/UNAIDS_FactSheet_en. pdf

6. UNAIDS. AIDSinfo. Country factsheets. Cameroon 2018. 2019; Available from: https://aidsinfo.unaids.org/

7. Sam-Agudu NA, Folayan MO, Ezeanolue EE. Seeking wider access to HIV testing for adolescents in sub-Saharan Africa. Pediatr Res. 2016;79(6):838–45.

8. Msellati P, Ateba Ndongo F, Hejoaka F, Nacro B. [Impediments to HIV testing in HIV-infected children and teenagers in Africa: look for them where they are!]. Med Sante Trop. 2016 Mar;26(1):10–4.

9. Eba PM, Lim H. Reviewing independent access to HIV testing, counselling and treatment for adolescents in HIV-specific laws in sub-Saharan Africa: implications for the HIV response. J Int AIDS Soc. 2017 11;20(1):21456.

10. PEPFAR Blueprint.pdf [Internet]. [cited 2020 May 17]. Available from: https://www.avac.org/sites/default/files/resource-files/PEPFAR%20Blueprint.pdf

11. Kellerman SE, Ahmed S, Feeley-Summerl T, Jay J, Kim MH, Koumans E, et al. Beyond PMTCT: Keeping HIV Exposed and Positive children healthy and alive. AIDS Lond Engl. 2013 Nov;27(0 2):S225–33.

12. PMTCT HIV Dept brief Oct 07.pdf [Internet]. [cited 2020 May 17]. Available from: https://www.who.int/hiv/pub/toolkits/PMTCT%20HIV%20Dept%20brief%20Oct%2007.pdf

13. World Health Organization, Joint United Nations Programme on HIV/AIDS. Guidance on provider-initiated HIV testing and counselling in health facilities. Geneva: World Health Organization; 2007.

14. Ahmed S, Kim MH, Sugandhi N, Phelps BR, Sabelli R, Diallo MO, et al. Beyond early infant diagnosis: case finding strategies for identification of HIV-infected infants and children. AIDS Lond Engl. 2013 Nov;27 Suppl 2:S235–245.

15. Leon N, Lewin S, Mathews C. Implementing a provider-initiated testing and counselling (PITC) intervention in Cape town, South Africa: a process evaluation using the normalisation process model. Implement Sci IS. 2013 Aug 26;8:97.

16. Davies M-A, Kalk E. Provider-Initiated HIV Testing and Counselling for Children. PLoS Med. 2014 May 27;11(5):e1001650.

17. Lallemant M, Chang S, Cohen R, Pecoul B. Pediatric HIV — A Neglected Disease? N Engl J Med. 2011 Aug 18;365(7):581–3.

18. WHO. Consolidated guidelines on HIV testing services. Geneva; Switzerland; 2015. Available from: http://www.who.int/hiv/pub/guidelines/hiv-testing-services/en/. Accessed on 15.05.2020.

19. Yumo HA, Kuaban C, Ajeh RA, Nji AM, Nash D, Kathryn A, et al. Active case finding: comparison of the acceptability, feasibility and effectiveness of targeted versus blanket provider-initiated-testing and counseling of HIV among children and adolescents in Cameroon. BMC Pediatr. 2018 25;18(1):309.

20. WHO. Research for an AIDS Free Generation: A global research agenda for paediatric HIV. Geneva, Switzerland; 2017. Available from: http://www.who.int/hiv/pub/toolkits/cipher-research-paediatric-hiv/en/

21. Ciaranello AL, Park J-E, Ramirez-Avila L, Freedberg KA, Walensky RP, Leroy V. Early infant HIV-1 diagnosis programs in resource-limited settings: opportunities for improved outcomes and more cost-effective interventions. BMC Med. 2011 May 20;9:59.

22. Anaba UC, Sam-Agudu NA, Ramadhani HO, Torbunde N, Abimiku A, Dakum P, et al. Missed opportunities for early infant diagnosis of HIV in rural North-Central Nigeria: A cascade analysis from the INSPIRE MoMent study. PLoS ONE [Internet]. 2019 Jul 31 [cited 2020 May 16];14(7). Available from: https://www.ncbi.nlm.nih.gov/pmc/articles/PMC6668908/

23. Arora DR, Maheshwari M, Arora B. Rapid Point-of-Care Testing for Detection of HIV and Clinical Monitoring. ISRN AIDS [Internet]. 2013 May 23 [cited 2020 May 15];2013. Available from: https://www.ncbi.nlm.nih.gov/pmc/articles/PMC3767371/

24. Spooner E, Govender K, Reddy T, Ramjee G, Mbadi N, Singh S, et al. Point-of-care HIV testing best practice for early infant diagnosis: an implementation study. BMC Public Health. 2019 ;19(1):731.

25. Bianchi F, Cohn J, Sacks E, Bailey R, Lemaire J-F, Machekano R, et al. Evaluation of a routine point-of-care intervention for early infant diagnosis of HIV: an observational study in eight African countries. Lancet HIV. 2019;6(6):e373–81.

26. UNAIDS. National HIV estimates files. Available from: https://www.unaids.org/en/dataanalysis/datatools/spectrum-epp. Accessed on 15.05.2020.

27. UNAIDS. Fact Sheet 2016. Global Statistics 2015. Available from: www.unaids.org/sites/default/files/media_asset/20150901_FactSheet_2015_en.pdf. Accessed on 15.05.2020.

28. Marwa R, Anaeli A. Perceived Barriers Toward Provider-Initiated HIV Testing and Counseling (PITC) in Pediatric Clinics: A Qualitative Study Involving Two Regional Hospitals in Dar-Es-Salaam, Tanzania. HIV/AIDS - Research and Palliative Care. 2020;12.

29. Kranzer K, Meghji J, Bandason T, Dauya E, Mungofa S, Busza J, et al. Barriers to Provider-Initiated Testing and Counselling for Children in a High HIV Prevalence Setting: A Mixed Methods Study. PLoS Med. 2014;11(5).

30. World Health Organization and UNICEF. Policy requirements for HIV testing and counselling of infants and young children in health facilities. 2010; Available from: https://apps.who.int/iris/bitstream/handle/10665/44276/9789241599092_eng.pdf. Accessed on 15.05.2020.

31. Wagner AD, Wachira CM, Njuguna IN, Maleche-Obimbo E, Sherr K, Inwani IW, et al. Active referral of children of HIV-positive adults reveals high prevalence of undiagnosed HIV. J Acquir Immune Defic Syndr 1999. 2016 Dec 15;73(5):e83–9.

32. Ahmed S, Sabelli RA, Simon K, Rosenberg NE, Kavuta E, Harawa M, et al. Index case finding facilitates identification and linkage to care of children and young persons living with HIV/AIDS in Malawi. Trop Med Int Health TM IH. 2017;22(8):1021–9.

33. Penda CI, Moukoko CEE, Koum DK, Fokam J, Meyong CAZ, Talla S, et al. Feasibility and utility of active case finding of HIV-infected children and adolescents by provider-initiated testing and counselling: evidence from the Laquintinie hospital in Douala, Cameroon. BMC Pediatr. 2018 03;18(1):259.

34. Yumo HA, Ajeh RA, Beissner M, Ndenkeh JN, Sieleunou I, Jordan MR, et al. Effectiveness of symptom-based diagnostic HIV testing versus targeted and blanket provider-initiated testing and counseling among children and adolescents in Cameroon. PLoS ONE. 2019;14(5).

